# The Role of Vaccine Status Homophily in the COVID-19 Pandemic: A Cross-Sectional Survey with Modeling

**DOI:** 10.1101/2023.06.06.23291056

**Authors:** Elisha B. Are, Kiffer G. Card, Caroline Colijn

## Abstract

**Background:** Vaccine homophily describes non-heterogeneous vaccine uptake within contact networks. This study was performed to determine observable patterns of vaccine homophily, associations between vaccine homophily, self-reported vaccination, COVID-19 prevention behaviours, contact network size, and self-reported COVID-19, as well as the impact of vaccine homophily on disease transmission within and between vaccination groups under conditions of high and low vaccine efficacy.

**Methods:** Residents of British Columbia, Canada, aged ≥16 years, were recruited via online advertisements between February and March 2022, and provided information about vaccination status, perceived vaccination status of household and non-household contacts, compliance with COVID-19 prevention guidelines, and history of COVID-19. A deterministic mathematical model was used to assess transmission dynamics between vaccine status groups under conditions of high and low vaccine efficacy.

**Results:** Vaccine homophily was observed among the 1304 respondents, but was lower among those with fewer doses (*p*<0.0001). Unvaccinated individuals had larger contact networks (*p*<0.0001), were more likely to report prior COVID-19 (*p*<0.0001), and reported lower compliance with COVID-19 prevention guidelines (*p*<0.0001). Mathematical modelling showed that vaccine homophily plays a considerable role in epidemic growth under conditions of high and low vaccine efficacy. Further, vaccine homophily contributes to a high force of infection among unvaccinated individuals under conditions of high vaccine efficacy, as well as elevated force of infection from unvaccinated to vaccinated individuals under conditions of low vaccine efficacy.

**Interpretation:** The uneven uptake of COVID-19 vaccines and the nature of the contact network in the population play important roles in shaping COVID-19 transmission dynamics.

## Introduction

COVID-19 is a respiratory illness caused by severe acute respiratory syndrome coronavirus 2 (SARS-CoV-2), which is transmitted predominantly via aerosols and droplets (1). In high-income countries, the general population case fatality rate of COVID-19 is sufficiently high to necessitate widespread public health interventions and targeted protections for vulnerable populations, such as seniors and people who are immunocompromised (2).

Fortunately, several safe and effective vaccines are available that can prevent severe COVID-19 and reduce mortality risk, although they have lower effectiveness against transmission than initially hoped (3). At the individual level, the effectiveness of these vaccines wanes over time, and is subject to immune escape (4). At the population level, the effectiveness of these vaccines is also dependent on their uptake within and across geographic regions and social networks (5,6). Of course, vaccine uptake is heterogeneous within any given population, and this heterogeneity may create disproportionate risk for SARS-CoV-2 transmission within and across communities.

Vaccine hesitancy is an important factor shaping vaccine uptake (7). A 2014 systematic review documented a range of factors that influence vaccine hesitancy, including contextual influences (e.g., politics, government, religion, geographic patterns, media); individual and social group influences (e.g., beliefs, attitudes, knowledge, trust in healthcare systems and providers); and vaccine-specific issues (e.g., mode of administration and delivery, vaccination schedules, risk vs. benefit) (8). The results showed that vaccine status tends to cluster with sociodemographic characteristics, such as age, socioeconomic status, race/ethnicity, and political orientation (8–10).

Homophily is a principle in sociology and mathematical modelling that describes the clustering of individual-level characteristics, such as vaccination status, with social networks (11). Kadelka and McCombs (12) suggested that vaccine homophily may impact COVID-19 vaccine effectiveness given the potential for uneven vaccination uptake. Modelling studies have explored the impact of homophily in a range of context, and its impact on transmission dynamics is well documented (13,14). For instance, a fairly recent modelling study argued that the mixing of vaccinated and unvaccinated groups contributes to considerable risk of infection at the population level (15). However, these previous studies were not based on descriptive data regarding vaccine homophily. Broadly, empirical research related to vaccine homophily in the context of the COVID-19 pandemic has been very limited. Therefore, it is important to describe COVID-19 vaccine homophily and its relationship to vaccination status to gain an improved understanding of COVID-19 transmission (16–19). This paper makes important contributions by connecting vaccination and contact heterogeneity, which are two crucial determinants of transmission dynamics. Furthermore, it assessed their impacts under low and high vaccination efficacy scenarios. Moreover, our model is fully informed and driven by the survey data.

The present study was performed to characterize observable patterns of vaccine homophily and examine associations between vaccine homophily, self-reported vaccination, COVID-19 prevention behaviours, contact network size, and self-reported COVID-19 infection; and to determine the impact of vaccine homophily on COVID-19 transmission within and between vaccination groups under conditions of high and low vaccine efficacy.

## Methods

### Participant Recruitment

Participants were recruited using paid Facebook advertisements (Figure 1) between February 16, 2022, and March 3, 2022, a period during which the average number of new COVID-19 cases in British Columbia was declining (7-Day Rolling Average: 865 on February 16, 487 on March 3) and the province continued to experience high numbers of Omicron variant infections (20).

**Figure 1.**
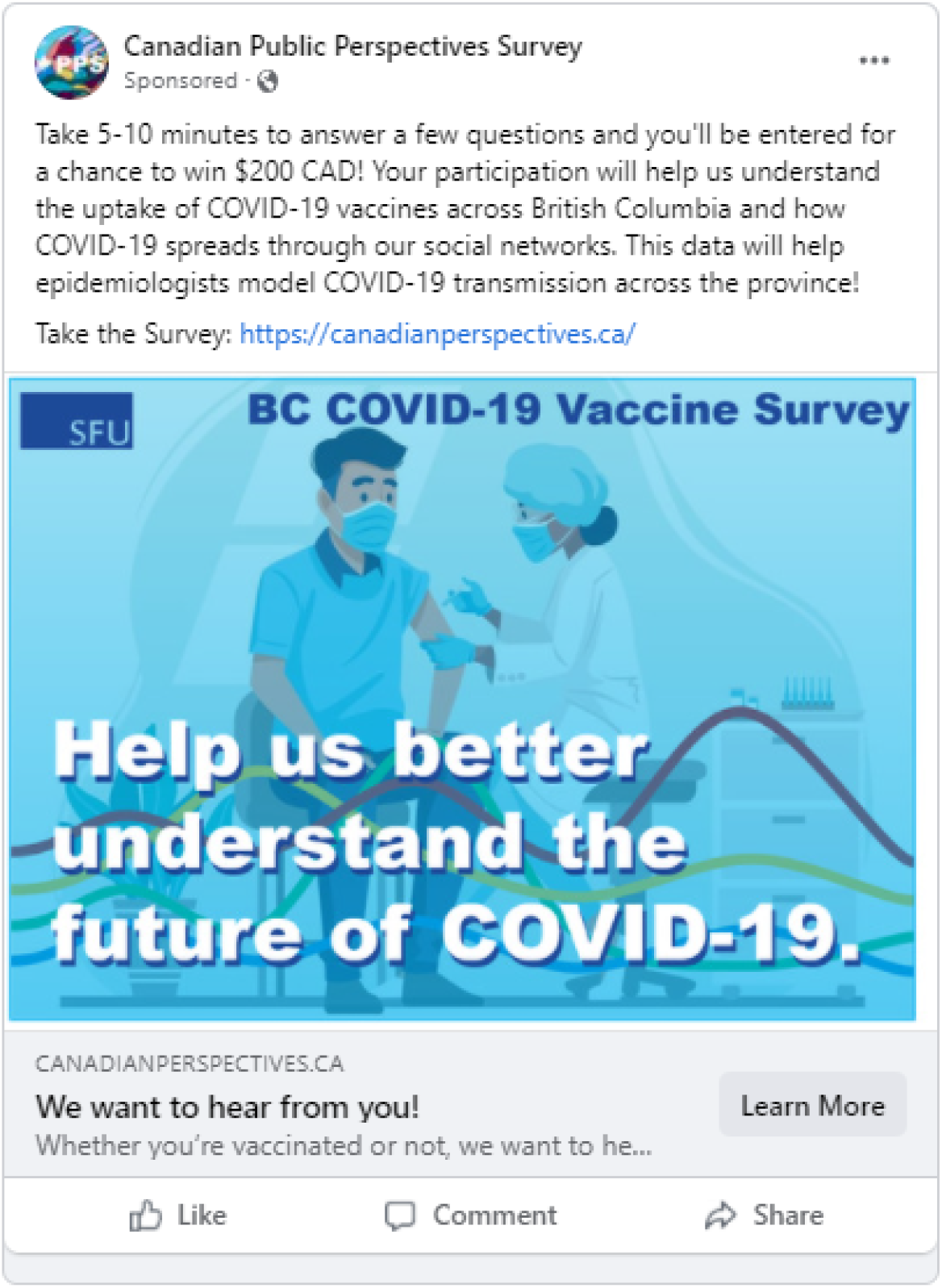
Facebook Advertisement Used for Participant Recruitment.

### Data Collection

After providing informed consent, potential participants recruited via Facebook advertisements were screened for eligibility. The eligibility criteria restricted participation to individuals aged 16 years or older living in British Columbia, Canada. Participants completed an online survey delivered in English using the Qualtrics platform, which assessed participants’ history of COVID-19, the extent to which they were following provincial mandates and guidelines for COVID-19 prevention, and how many COVID-19 vaccine doses they had received. Participants also reported on the perceived COVID-19 history of their regular contacts, the perceived level of compliance to COVID-19 prevention guidelines and mandates among regular contacts, the vaccination status of their household and non-household contacts, and the number of household and non-household contacts with whom they had recent contacts. Supplemental Table S1 provides an overview of how these variables were measured by providing the question text and response options.

Additionally, the following demographic data of the participants were collected: age (numerical), gender (Male; Female; Non-binary), ethnicity (African, Caribbean, or Black; Arab or West Asia; East Asian; Indigenous; Latin American; South Asian; Southeast Asian; White; Other), education level (Some high school; High school diploma or equivalent; Some college or trades training; Some university; College or trades certificate or diploma; University degree or higher), annual household income ($0 to $150,000 or higher), and whether participants were born in Canada (Yes; No, moved to Canada in the last 5 years; No, moved to Canada more than 5 years ago). Postal code was also assessed and was used to assign participants to one of the 5 regional health authorities in British Columbia.

## Data Analysis

### Aim 1. Characterization of Vaccine Homophily and its Relationship to COVID-19 Transmission Dynamics

To characterize observable patterns of vaccine homophily and examine associations between vaccine homophily, self-reported vaccination status, COVID-19 prevention behaviours, contact network size, and self-reported COVID-19 infection, descriptive analyses of survey responses were conducted in R version 4.1.3. (21). Data were cleaned using the Tidyverse collection of R packages (22). As a preliminary step, participants with missing data on demographic-variables (i.e., age, gender, ethnicity, income, education level, immigration status, and health authority) or poor-quality responses (i.e., those in which incongruent responses were provided across questions, indicating imprecise answering) were removed from the analysis. Removal of data with missing demographic variables was done because our sample weighting procedure was not tolerant of missing data (23). The remaining observations were weighted by weighting variables using iterative proportional fitting raking estimation, which is a well-established approach for multivariable weighting when only the marginal proportions for each variable are known (24–26). Raking estimation was implemented using the anesrake package (27) with marginal proportions for each weighting variable derived from the 2016 Canadian Census Profile for British Columbia (28). The survey package was used to generate weighted descriptive statistics (29). Weighted descriptive data were plotted using the questionr and ggplot2 packages (30,31). For descriptive statistics, all observations included in the weighted sample were included allowing for us to maximize the information provided by participants without removing them due to non-response (e.g., early survey drop off, refusal to answer). All variables had less than 5% missingness.

To understand clustering between risk factors for COVID-19 and participant’s self-reported vaccination status, the chi-square and Kruskal–Wallis tests were used to compare participants with 0, 1, 2, or 3 or more doses with regard to their personal history of COVID-19; personal compliance with provincial COVID-19 prevention guidelines; perceived prevalence of past COVID-19 diagnoses among regular contacts; perceived prevalence of vaccination among regular contacts; perceived prevalence of very close adherence to provincial COVID-19 prevention practices among regular contacts; household, non-household, and overall number of contacts; vaccine prevalence among household, non-household, and overall contacts; and overall proportion of contacts with the same vaccination status (i.e., vaccine homophily).

- ***Average Number of Doses Among Contacts.*** The average numbers of doses among household, non-household, and overall contacts were calculated using self-reported estimates of the number of doses that participants believed each of their contacts had received.
- ***Proportion of Contacts with* ≥*1 Dose.*** The proportion of contacts with ≥1 dose was calculated using self-reported data on the number of doses that participants believed each of their contacts had received.
- ***External-Internal Homophily Index Score.*** The external-internal (E-I) homophily index was calculated as the overall proportion of contacts with the same vaccination status (i.e., vaccine homophily) as the participant. Values were calculated for household, non-household, and overall contacts.
- ***Prevalence-Adjusted Homophily Score.*** As homophily is a function of the general prevalence of each participant’s vaccination status, we calculated the prevalence-adjusted homophily (PAH) score, which is a simple statistic that measures whether the homophily in a participant’s vaccination status was above or below the expected level. The PAH score was calculated by subtracting the fraction of the entire sample with the same vaccination status as the participant from the fraction of each participant’s contact network with the same vaccination status. Values for household, non-household, and overall contact networks were calculated as follows:

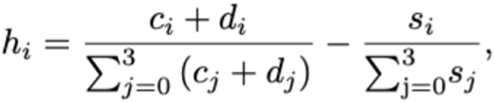

where *h*_*i*_ is the PAH score of an individual who has received *i* doses, *c*_*i*_ is the number of people in the contact network of an individual with *i* doses who have also received *i* doses, *d_i_* is the number of people in the household of an individual with *i* doses who have received *i* doses themselves, and *s*_*i*_ is the number of individuals in the sample who have received *i* vaccine doses. Each participant therefore has their own *h*_*i*_ value. The mean PAH score was calculated for each vaccination group.

This resulted in a score in which positive and negative values represented higher and lower than anticipated degree of vaccine homophily, respectively. This was analyzed descriptively to confirm that the relationship of the E-I homophily index value to personal number of vaccine doses was not merely a function of the level of vaccination in the sample (and reflecting the true vaccination prevalence in the population).

- ***Blau’s Heterogeneity Index Score.*** For each participant, we also calculated the diversity of vaccination statuses in their social network using Blau’s heterogeneity index, calculated as 1 minus the sum (over the numbers of doses, *k*) of the squared fraction of the participant’s contact networks with *k* doses (*p_k_ ^2^)*:

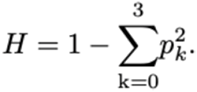

Blau’s heterogeneity index scores were calculated for the number of doses (*k*=0, 1, 2, 3) for each participant’s overall contact network and dichotomized vaccination status (i.e., *k*≥1 dose vs. <1 dose) for each participant’s household, non-household, and overall contact networks.

All homophily estimates and diversity estimates were calculated across each level of vaccination, and associations with self-reported vaccination, COVID-19 prevention behaviours, contact network size, and self-reported COVID-19 were tested using the Kruskal–Wallis H test. Associations between continuous measures were assessed using Spearman’s rank correlation test.

### Aim 2. Demonstration of the Impact of Vaccine Homophily on COVID-19 Transmission

To demonstrate the impact of vaccine homophily on COVID-19 transmission within and between vaccination groups under conditions of high and low vaccine efficacy, we developed a deterministic model that accounts for heterogeneity in contact patterns to assess the dynamic impact of vaccine homophily on COVID-19 transmission in British Columbia. We analyzed the effects of vaccine homophily under two broad scenarios with low and high vaccine efficacy against infection.

#### Model assumptions

The present model was designed to illustrate the impact of vaccine homophily under conditions of low and high vaccine efficacy. The model population was stratified according to the number of COVID-19 vaccine doses received. Interactions within and between groups interactions occur with different contact rates and preferences, reflecting the extent to which individuals contact others with their own vaccine status vs others. Vaccine effectiveness against infection is not 100%, so breakthrough infections can occur in all groups in the model, with a rate depending on exposure and on the number of doses and vaccine efficacy. Immunity wanes at a constant rate; here, this means immunity against infection (disease was not modelled explicitly, because the focus is on the transmission dynamics). Finally, we focused on a short time period during which vaccination levels in the population were maintained.

#### Model Equation

The model equations are:

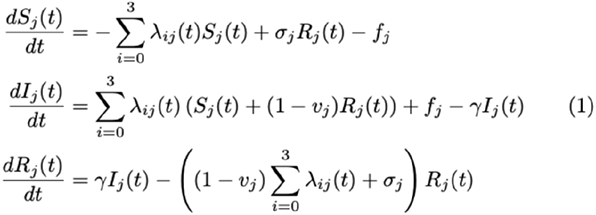

where *i*, *j* = 0, 1, 2, 3 represent the number of COVID-19 doses an individual has received.

Table 1 shows a description of the variables and parameters used in the model.

**Table 1.** Descriptions of Variables and Parameters

| Variables and parameters | Description and sources |
| --- | --- |
| $S_j(t)$ | Number of susceptible individuals |
| $I_j(t)$ | Number of infectious individuals |
| $R_j(t)$ | Number of recovered individuals |
| $\sigma_j$ | Waning rate per day for immunity against infection. Set at 1/(183 days) |
| $f_j$ | Importation rate (e.g., due to travel). Set at 150 infections per day for those with $\geq 2$ doses and 0 for those with $< 2$ doses. (32). Assuming that travel restrictions are effective. |
| $v_j$ | Strength of short-term protection from reinfection. At baseline: $v_0 = 0.35, v_1 = 0.65, v_2 = 0.68, v_3 = 0.83$ (Assumed) |
| $\gamma$ | Recovery rate per day. Set at 1/ (4 days) (33) |
| $\beta$ | Probability of infection given contact: 0.2 (Fitted) |
| $p_{ji}$ | Proportion of contacts of those with vaccination status $j$ that are of vaccination status $i$ (Estimated form survey data) |
| $c_j$ | The total number of contacts per day made by individuals with $j$ doses (Estimated form survey data) |
| $l_j$ | The level to which those with $j$ doses comply with physical distancing measures (Estimated form survey data) |

#### Force of infection

The force of infection λ(*t*) was defined as: (number of contacts per unit time) × (probability of disease transmission per contact) × (proportion of contacts that are infected). We used the following expression to model the force of infection:

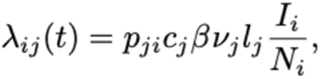

where λ_*ij*_ (*t*) is the force of infection for transmitting infection from individuals with vaccination status *i* to those with vaccination status *j*, *p*_*ji*_ is the proportion of contacts of those with vaccination status *j* that are of vaccination status *i*, *c*_*j*_ is the total number of contacts per day made by individuals with *j* doses, β is the probability of infection given contact, ν_*j*_ is vaccine efficacy against infection for individuals with *j* doses, *l*_*j*_ is the level to which those with *j* doses comply with physical distancing measures, *I*_*i*_ is the number of infected individuals who have had *i* doses, and *N*_*i*_ is the total number of people with *i* doses. Parameter values were extracted from the Facebook survey data.

#### Model validation

We matched model output to reported cases of COVID-19 during the survey period from February 16 to March 3, 2022. We accounted for underreporting of cases by assuming a constant underascertainment probability during the study period. The model yielded a good fit to the data and provided reasonable initial conditions for subsequent model prediction. The model fit to data is shown in Fig. S1.

#### Model scenarios

We analyzed the impact of vaccine homophily on COVID-19 transmission dynamics under two broad scenarios. First, we assumed that vaccine efficacy in preventing infection is relatively high, representing conditions where a reasonable proportion of the population has recently received a booster vaccination. This corresponds to the situation prior to the emergence of the Omicron variant, which showed substantial escape from immunity against infection, or future scenarios where more effective vaccines are available and have been widely used. Second, we modelled a scenario with low vaccine efficacy, representing time periods where immunity has waned significantly or when the dominant variant shows low sensitivity to vaccine protection. We further considered each of the above scenarios with and without homophily. For the former, we used contact-related parameter (*p*_*ji*_, *c*_*j*_, *l*_*j*_) values estimated from the survey data, while in the latter, we calculated a weighted average for each of the parameters to eliminate the impact of vaccine homophily. That is, for the without homophily scenario, the total number of contacts (*c*_*j*_) for each vaccination group and the proportion of contacts individuals make with those in their group and everyone else, as well as the level of adherence to physical distancing measures, are the same for each group regardless of vaccination status.

### Ethics Review

The study protocol was approved by the Research Ethics Board of Simon Fraser University (Protocol #30000753). All participants provided informed consent before completing the survey.

## Results

### Aim 1. Characterization of Vaccine Homophily and its Relationship to COVID-19 Transmission Dynamics

Facebook and Instagram advertisements were displayed to 266,894 users. A total of 3659 participants initiated the survey and provided informed consent to participate in the study. After exclusion of responses that were of poor quality or had missing data, the final analytical sample size was 1185.

The unweighted sample was disproportionately White (86.9%), female (58.1%), had higher income (≥$90,000, 58.7%), and had been born in Canada (82.8%) (**Table 2**). Statistical weights were used to align these factors with the population distribution based on the 2016 Canadian Census Profile for British Columbia.

**Table 2.**
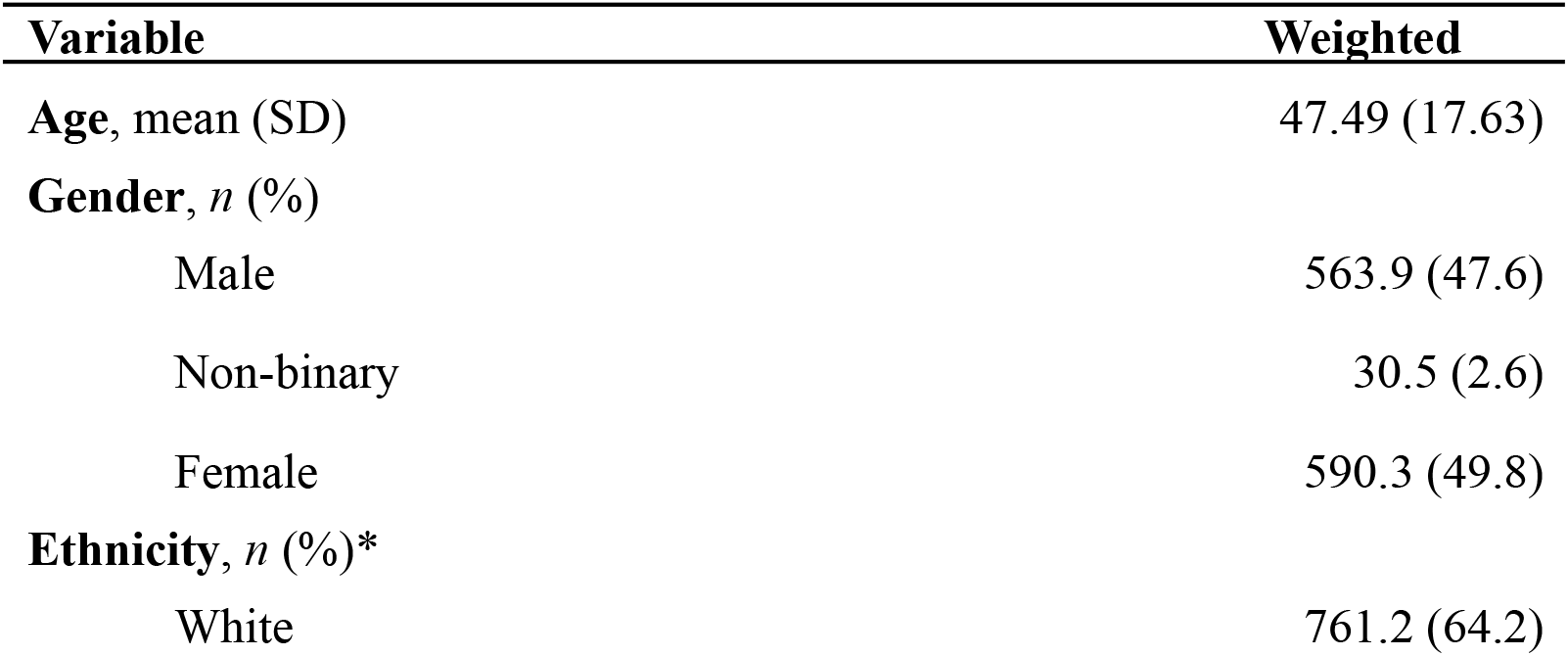
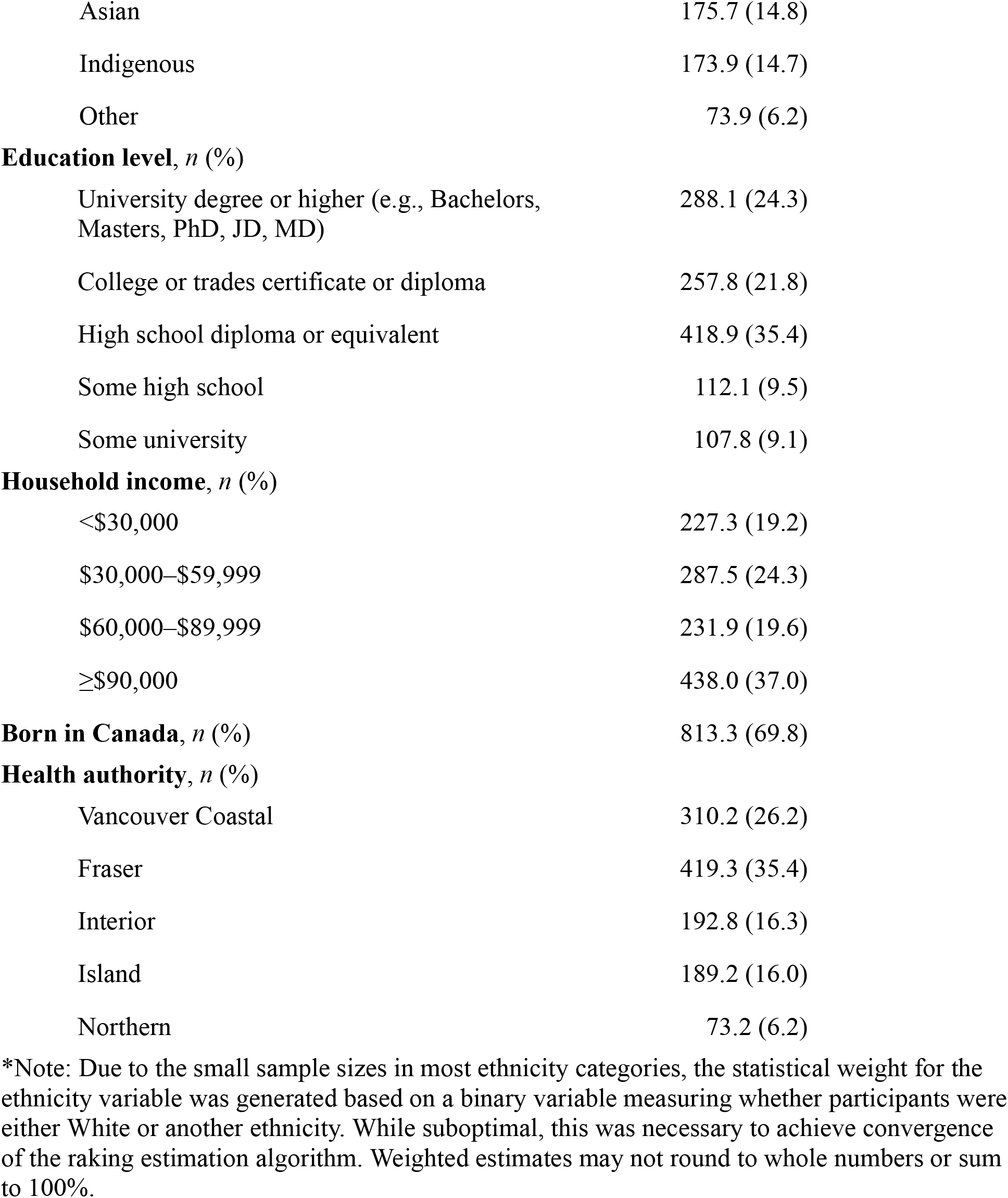
Characteristics of the Study Population, Weighted for BC Population Characteristics based on 2016 Census Profile

**Table 3** presents additional descriptive statistics about self-reported COVID-19 diagnosis history and self-reported compliance with provincial COVID-19 prevention guidelines, stratified by self-reported vaccination status. In summary, participants who had received more doses of the COVID-19 vaccine were less likely to report a previous COVID-19 diagnosis (*p*<0.0001) and were more likely to report higher compliance with provincial COVID-19 prevention guidelines (*p*<0.0001).

**Table 3.** Personal Indicators of COVID-19 Risk, Weighted

|  | 0 Doses | 1 Dose | 2 Doses | ≥3 Doses |
| --- | --- | --- | --- | --- |
|  | n =<br>234.4 | n =<br>20.6 | n =<br>243.9 | n =<br>685.7 |
| **COVID-19 Diagnosis/Infection, *n* (%)** |  |  |  |  |
| No, and I do not think I have had COVID-19 | 76.3<br>(32.9) | 10.9<br>(59.6) | 73.2<br>(30.4) | 486.2<br>(74.0) |
| No, but I think I have had COVID-19. I just never received a test and/or diagnosis. | 86.6<br>(37.4) | 2.8<br>(15.1) | 92.0<br>(38.1) | 112.9<br>(17.2) |
| Yes, I have been diagnosed with COVID-19 | 69.0<br>(29.8) | 4.7<br>(25.3) | 75.9<br>(31.5) | 57.9<br>(8.8) |
| **Compliance with COVID-19 Guidelines, *n* (%)** |  |  |  |  |
| Not At All | 28.8<br>(12.3) | 1.0<br>(5.0) | 14.6<br>(6.0) | 0.4<br>(0.1) |
| Not Very Closely | 77.6<br>(33.1) | 4.5<br>(22.0) | 59.0<br>(24.3) | 13.3<br>(2.1) |
| Somewhat Closely | 97.1<br>(41.4) | 9.9<br>(48.3) | 94.6<br>(39.0) | 116.6<br>(18.0) |
| Very Closely | 30.9<br>(13.2) | 5.1<br>(24.8) | 74.4<br>(30.7) | 515.9<br>(79.8) |

*Note: Values may not round to whole numbers or sum to 100% due to missing observations on some variables and statistical weighting.*

**Table 4** presents descriptive statistics for participant-reported descriptions of their household and non-household contacts stratified according to self-reported vaccination status. Briefly, participants who had received more doses of the COVID-19 vaccine had networks with higher average numbers of doses, had a greater proportion of network contacts with at least one vaccine dose, had higher E-I homophily index scores, and had higher vaccine homophily. With regard to network vaccine heterogeneity, participants with fewer vaccine doses had more heterogeneous networks according to Blau’s heterogeneity index.

**Table 4.**
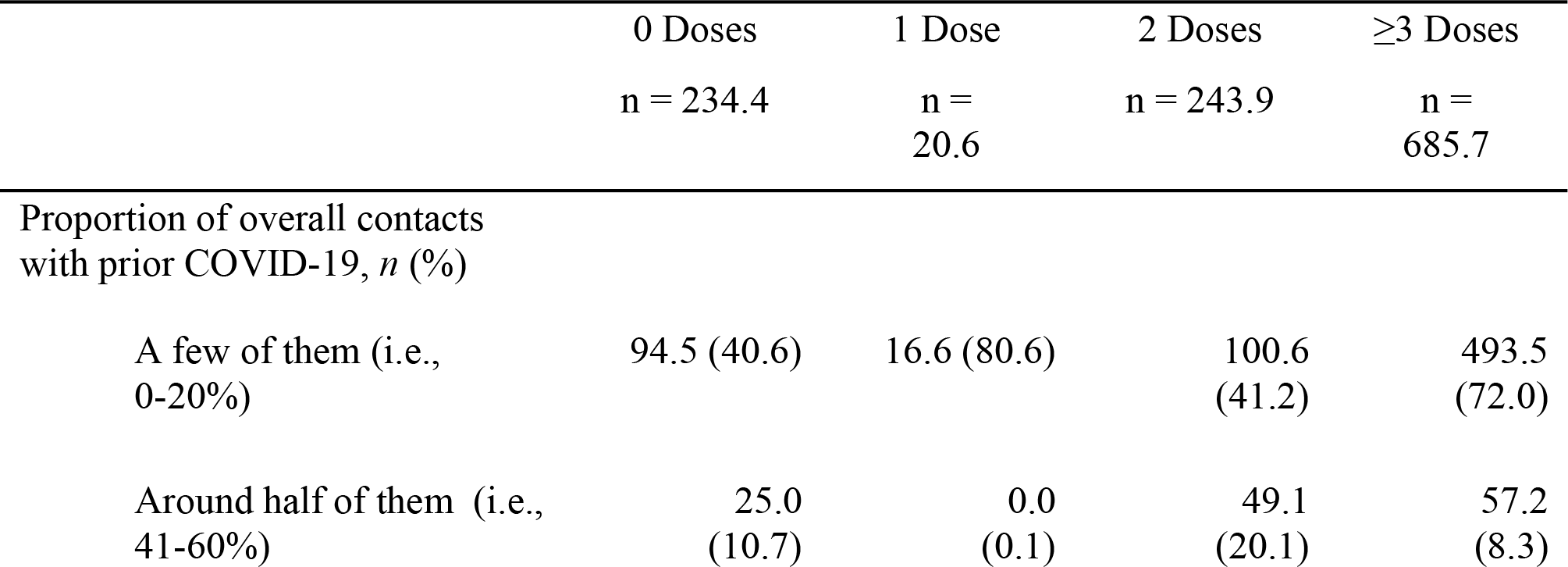
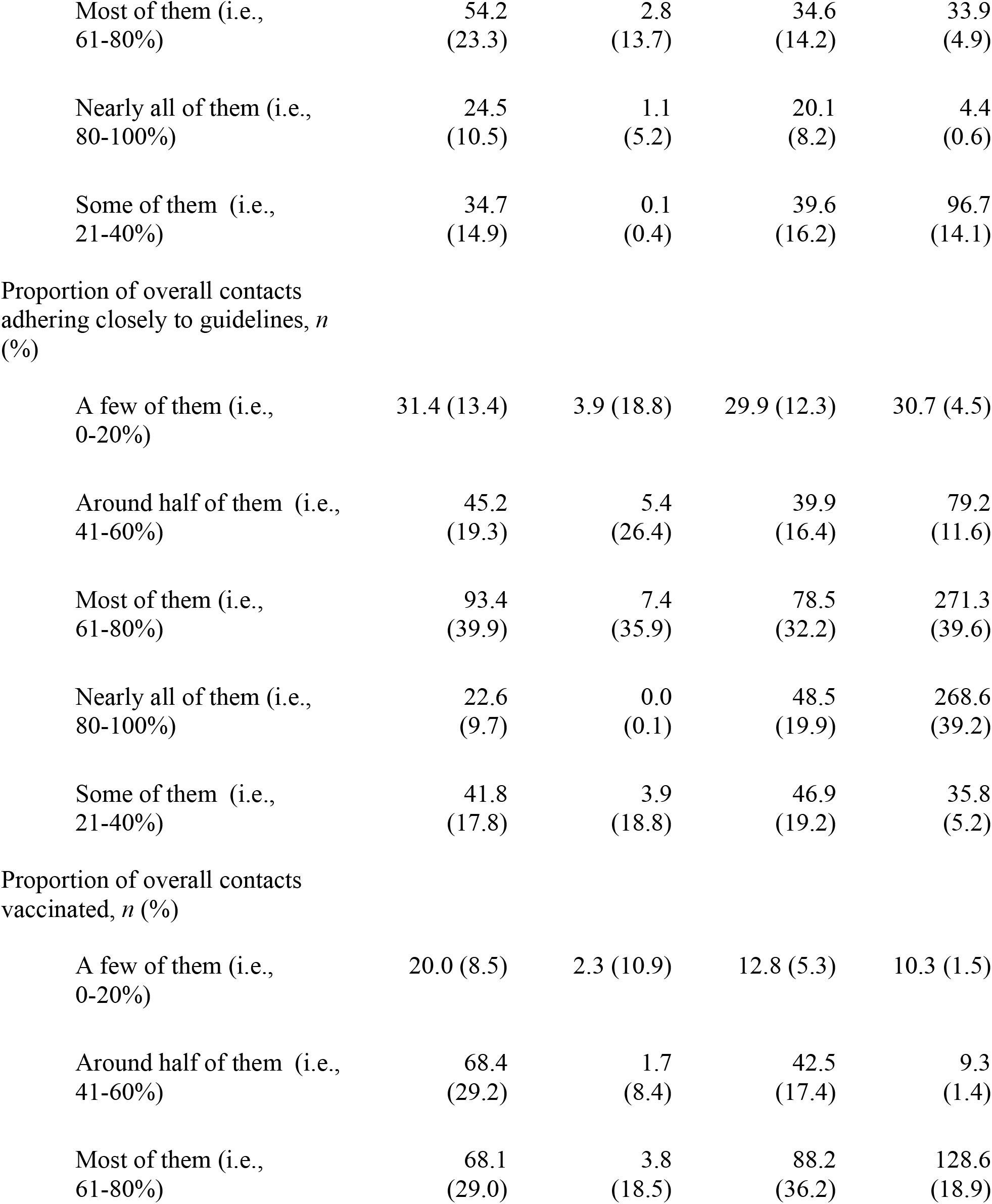
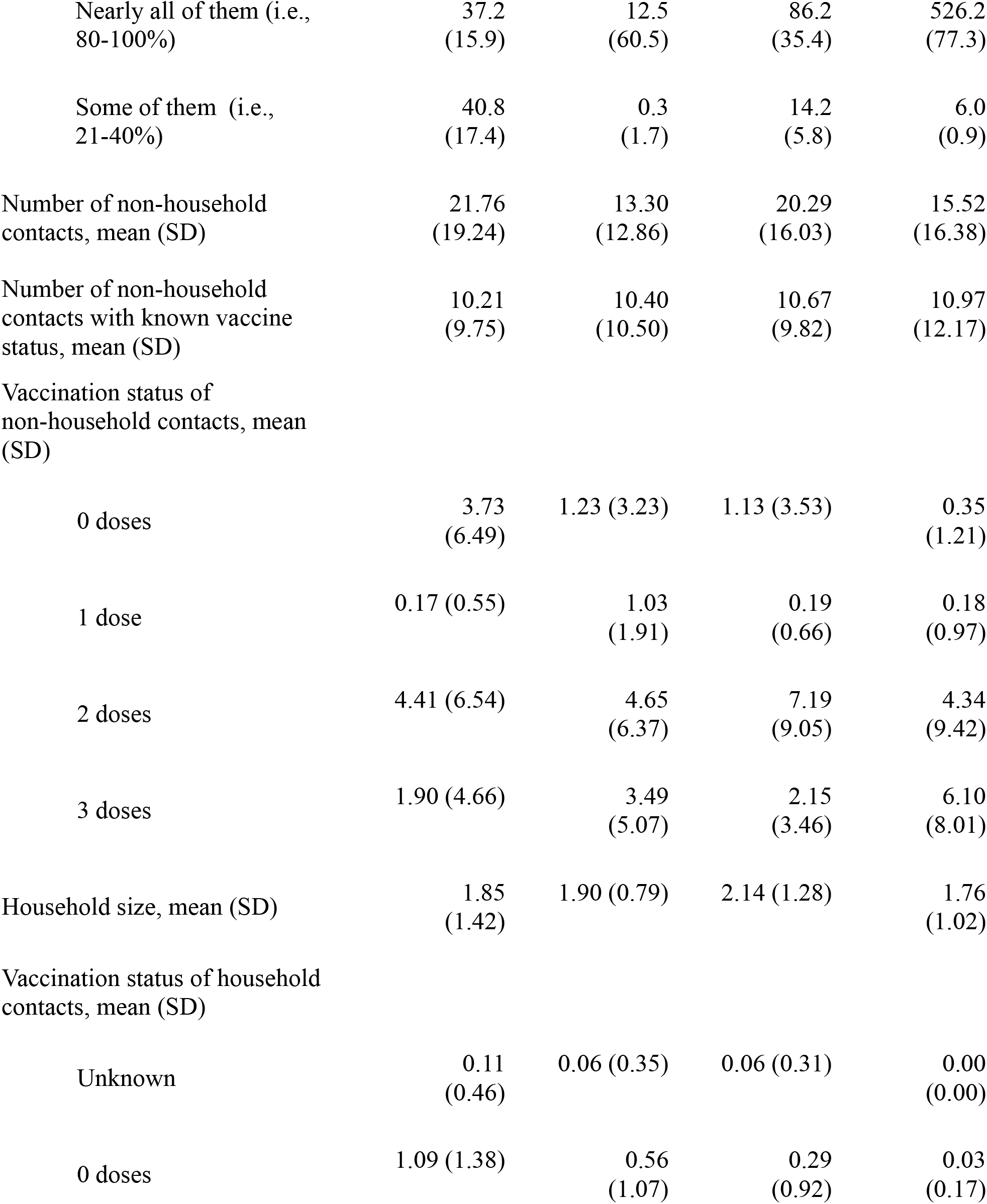
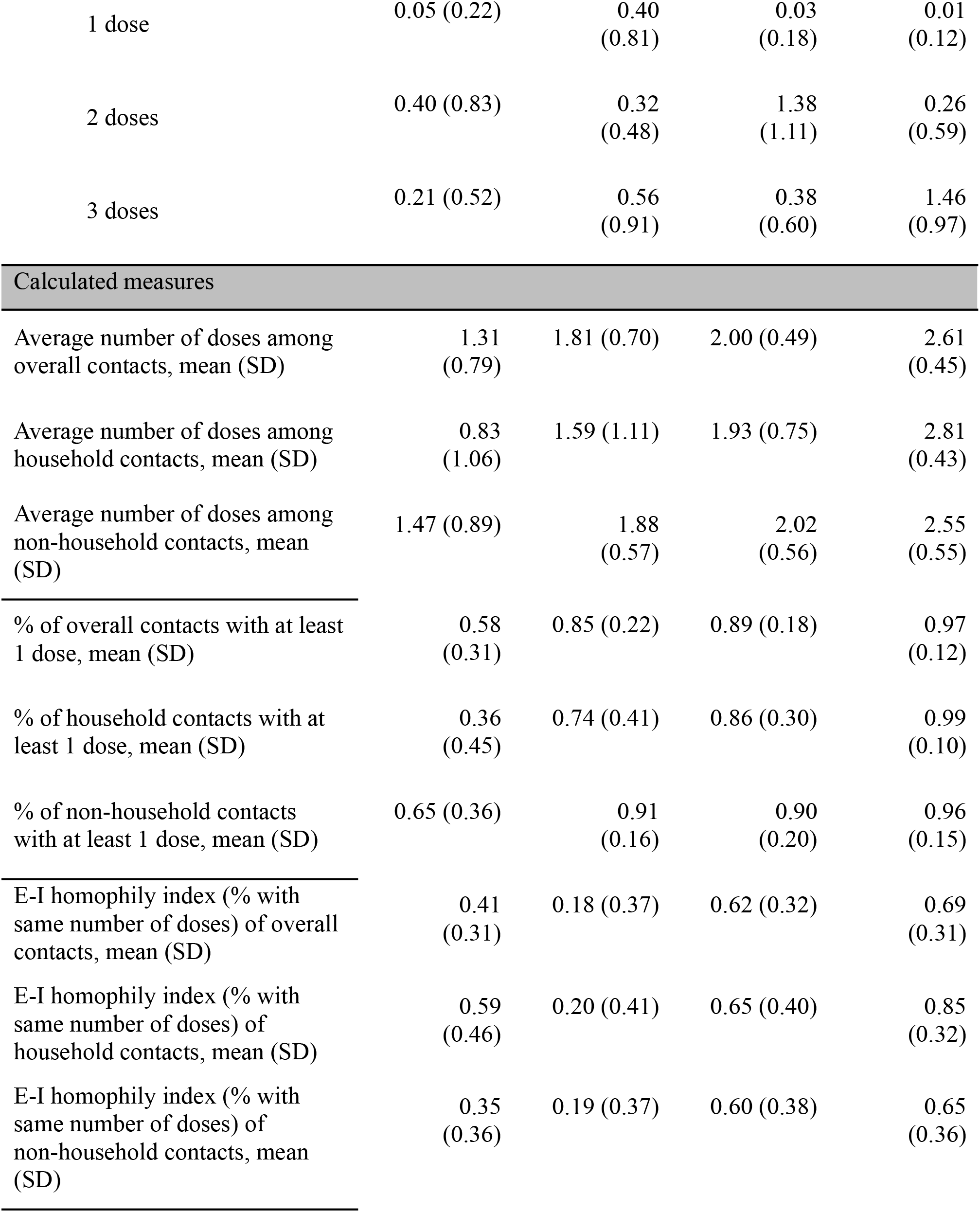
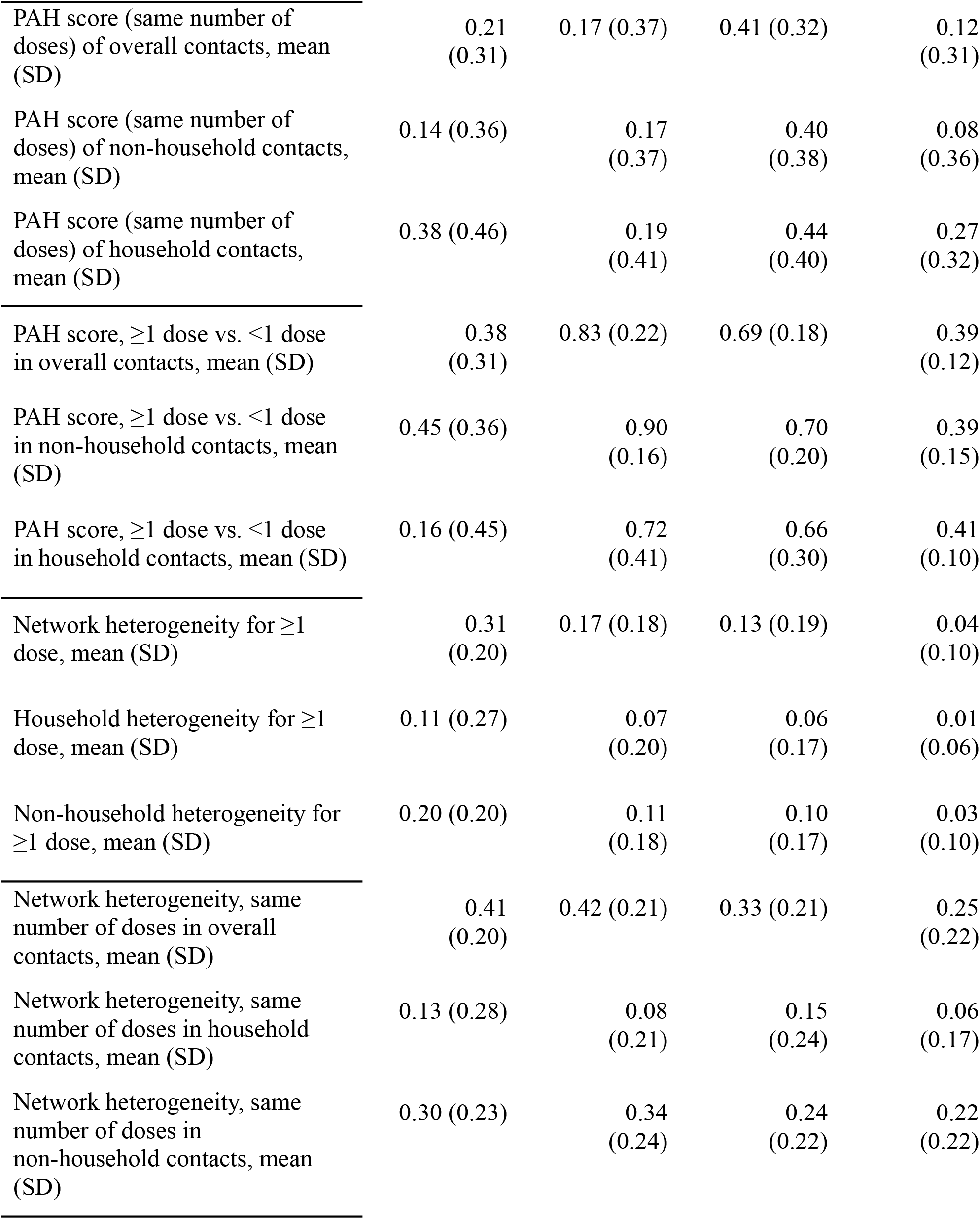
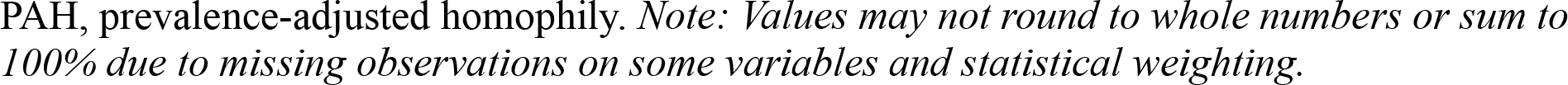
Social network indicators of COVID-19 risk

|  | 0 Doses<br>n = 234.4 | 1 Dose<br>n =<br>20.6 | 2 Doses<br>n = 243.9 | ≥3 Doses<br>n =<br>685.7 |
| --- | --- | --- | --- | --- |
| Proportion of overall contacts<br>with prior COVID-19, *n* (%) |  |  |  |  |
| A few of them (i.e.,<br>0-20%) | 94.5 (40.6) | 16.6 (80.6) | 100.6<br>(41.2) | 493.5<br>(72.0) |
| Around half of them (i.e.,<br>41-60%) | 25.0<br>(10.7) | 0.0<br>(0.1) | 49.1<br>(20.1) | 57.2<br>(8.3) |
| Most of them (i.e.,<br>61-80%) | 54.2<br>(23.3) | 2.8<br>(13.7) | 34.6<br>(14.2) | 33.9<br>(4.9) |
| Nearly all of them (i.e.,<br>80-100%) | 24.5<br>(10.5) | 1.1<br>(5.2) | 20.1<br>(8.2) | 4.4<br>(0.6) |
| Some of them (i.e.,<br>21-40%) | 34.7<br>(14.9) | 0.1<br>(0.4) | 39.6<br>(16.2) | 96.7<br>(14.1) |
| Proportion of overall contacts<br>adhering closely to guidelines, *n*<br>(%) |  |  |  |  |
| A few of them (i.e.,<br>0-20%) | 31.4 (13.4) | 3.9 (18.8) | 29.9 (12.3) | 30.7 (4.5) |
| Around half of them (i.e.,<br>41-60%) | 45.2<br>(19.3) | 5.4<br>(26.4) | 39.9<br>(16.4) | 79.2<br>(11.6) |
| Most of them (i.e.,<br>61-80%) | 93.4<br>(39.9) | 7.4<br>(35.9) | 78.5<br>(32.2) | 271.3<br>(39.6) |
| Nearly all of them (i.e.,<br>80-100%) | 22.6<br>(9.7) | 0.0<br>(0.1) | 48.5<br>(19.9) | 268.6<br>(39.2) |
| Some of them (i.e.,<br>21-40%) | 41.8<br>(17.8) | 3.9<br>(18.8) | 46.9<br>(19.2) | 35.8<br>(5.2) |
| Proportion of overall contacts<br>vaccinated, *n* (%) |  |  |  |  |
| A few of them (i.e.,<br>0-20%) | 20.0 (8.5) | 2.3 (10.9) | 12.8 (5.3) | 10.3 (1.5) |
| Around half of them (i.e.,<br>41-60%) | 68.4<br>(29.2) | 1.7<br>(8.4) | 42.5<br>(17.4) | 9.3<br>(1.4) |
| Most of them (i.e.,<br>61-80%) | 68.1<br>(29.0) | 3.8<br>(18.5) | 88.2<br>(36.2) | 128.6<br>(18.9) |
| Nearly all of them (i.e., 80-100%) | 37.2<br>(15.9) | 12.5<br>(60.5) | 86.2<br>(35.4) | 526.2<br>(77.3) |
| Some of them (i.e., 21-40%) | 40.8<br>(17.4) | 0.3<br>(1.7) | 14.2<br>(5.8) | 6.0<br>(0.9) |
| Number of non-household contacts, mean (SD) | 21.76<br>(19.24) | 13.30<br>(12.86) | 20.29<br>(16.03) | 15.52<br>(16.38) |
| Number of non-household contacts with known vaccine status, mean (SD) | 10.21<br>(9.75) | 10.40<br>(10.50) | 10.67<br>(9.82) | 10.97<br>(12.17) |
| Vaccination status of non-household contacts, mean (SD) |  |  |  |  |
| 0 doses | 3.73<br>(6.49) | 1.23 (3.23) | 1.13 (3.53) | 0.35<br>(1.21) |
| 1 dose | 0.17 (0.55) | 1.03<br>(1.91) | 0.19<br>(0.66) | 0.18<br>(0.97) |
| 2 doses | 4.41 (6.54) | 4.65<br>(6.37) | 7.19<br>(9.05) | 4.34<br>(9.42) |
| 3 doses | 1.90 (4.66) | 3.49<br>(5.07) | 2.15<br>(3.46) | 6.10<br>(8.01) |
| Household size, mean (SD) | 1.85<br>(1.42) | 1.90 (0.79) | 2.14 (1.28) | 1.76<br>(1.02) |
| Vaccination status of household contacts, mean (SD) |  |  |  |  |
| Unknown | 0.11<br>(0.46) | 0.06 (0.35) | 0.06 (0.31) | 0.00<br>(0.00) |
| 0 doses | 1.09 (1.38) | 0.56<br>(1.07) | 0.29<br>(0.92) | 0.03<br>(0.17) |
| 1 dose | 0.05 (0.22) | 0.40<br>(0.81) | 0.03<br>(0.18) | 0.01<br>(0.12) |
| 2 doses | 0.40 (0.83) | 0.32<br>(0.48) | 1.38<br>(1.11) | 0.26<br>(0.59) |
| 3 doses | 0.21 (0.52) | 0.56<br>(0.91) | 0.38<br>(0.60) | 1.46<br>(0.97) |
| Calculated measures |  |  |  |  |
| Average number of doses among overall contacts, mean (SD) | 1.31<br>(0.79) | 1.81 (0.70) | 2.00 (0.49) | 2.61<br>(0.45) |
| Average number of doses among household contacts, mean (SD) | 0.83<br>(1.06) | 1.59 (1.11) | 1.93 (0.75) | 2.81<br>(0.43) |
| Average number of doses among non-household contacts, mean (SD) | 1.47 (0.89) | 1.88<br>(0.57) | 2.02<br>(0.56) | 2.55<br>(0.55) |
| % of overall contacts with at least 1 dose, mean (SD) | 0.58<br>(0.31) | 0.85 (0.22) | 0.89 (0.18) | 0.97<br>(0.12) |
| % of household contacts with at least 1 dose, mean (SD) | 0.36<br>(0.45) | 0.74 (0.41) | 0.86 (0.30) | 0.99<br>(0.10) |
| % of non-household contacts with at least 1 dose, mean (SD) | 0.65 (0.36) | 0.91<br>(0.16) | 0.90<br>(0.20) | 0.96<br>(0.15) |
| E-I homophily index (% with same number of doses) of overall contacts, mean (SD) | 0.41<br>(0.31) | 0.18 (0.37) | 0.62 (0.32) | 0.69<br>(0.31) |
| E-I homophily index (% with same number of doses) of household contacts, mean (SD) | 0.59<br>(0.46) | 0.20 (0.41) | 0.65 (0.40) | 0.85<br>(0.32) |
| E-I homophily index (% with same number of doses) of non-household contacts, mean (SD) | 0.35<br>(0.36) | 0.19 (0.37) | 0.60 (0.38) | 0.65<br>(0.36) |
| PAH score (same number of doses) of overall contacts, mean (SD) | 0.21<br>(0.31) | 0.17 (0.37) | 0.41 (0.32) | 0.12<br>(0.31) |
| PAH score (same number of doses) of non-household contacts, mean (SD) | 0.14 (0.36) | 0.17<br>(0.37) | 0.40<br>(0.38) | 0.08<br>(0.36) |
| PAH score (same number of doses) of household contacts, mean (SD) | 0.38 (0.46) | 0.19<br>(0.41) | 0.44<br>(0.40) | 0.27<br>(0.32) |
| PAH score, $\geq 1$ dose vs. $< 1$ dose in overall contacts, mean (SD) | 0.38<br>(0.31) | 0.83 (0.22) | 0.69 (0.18) | 0.39<br>(0.12) |
| PAH score, $\geq 1$ dose vs. $< 1$ dose in non-household contacts, mean (SD) | 0.45 (0.36) | 0.90<br>(0.16) | 0.70<br>(0.20) | 0.39<br>(0.15) |
| PAH score, $\geq 1$ dose vs. $< 1$ dose in household contacts, mean (SD) | 0.16 (0.45) | 0.72<br>(0.41) | 0.66<br>(0.30) | 0.41<br>(0.10) |
| Network heterogeneity for $\geq 1$ dose, mean (SD) | 0.31<br>(0.20) | 0.17 (0.18) | 0.13 (0.19) | 0.04<br>(0.10) |
| Household heterogeneity for $\geq 1$ dose, mean (SD) | 0.11 (0.27) | 0.07<br>(0.20) | 0.06<br>(0.17) | 0.01<br>(0.06) |
| Non-household heterogeneity for $\geq 1$ dose, mean (SD) | 0.20 (0.20) | 0.11<br>(0.18) | 0.10<br>(0.17) | 0.03<br>(0.10) |
| Network heterogeneity, same number of doses in overall contacts, mean (SD) | 0.41<br>(0.20) | 0.42 (0.21) | 0.33 (0.21) | 0.25<br>(0.22) |
| Network heterogeneity, same number of doses in household contacts, mean (SD) | 0.13 (0.28) | 0.08<br>(0.21) | 0.15<br>(0.24) | 0.06<br>(0.17) |
| Network heterogeneity, same number of doses in non-household contacts, mean (SD) | 0.30 (0.23) | 0.34<br>(0.24) | 0.24<br>(0.22) | 0.22<br>(0.22) |

PAH, prevalence-adjusted homophily. *Note: Values may not round to whole numbers or sum to 100% due to missing observations on some variables and statistical weighting.*

**Figure 2** shows the density curves for the average number of doses among the overall contacts stratified according to self-reported vaccination status. This figure illustrates that the social network contacts of participants with more vaccine doses were also more likely to have received more doses, suggesting higher homophily. This association between the average number of vaccine doses among contacts and personal vaccination status was statistically significant (*p*<0.0001). All groups showed higher than expected homophily, as measured by PAH estimates. Notably, the lowest average vaccine homophily was observed for participants with only one dose of vaccine, followed by those who had received no doses. Those with two or three or more doses of the COVID-19 vaccine had similar vaccine homophily. Those with only one or no doses also had considerably higher heterogeneity in the average number of vaccine doses among their network contacts (i.e., their contacts included both vaccinated and unvaccinated individuals).

**Figure 2.**
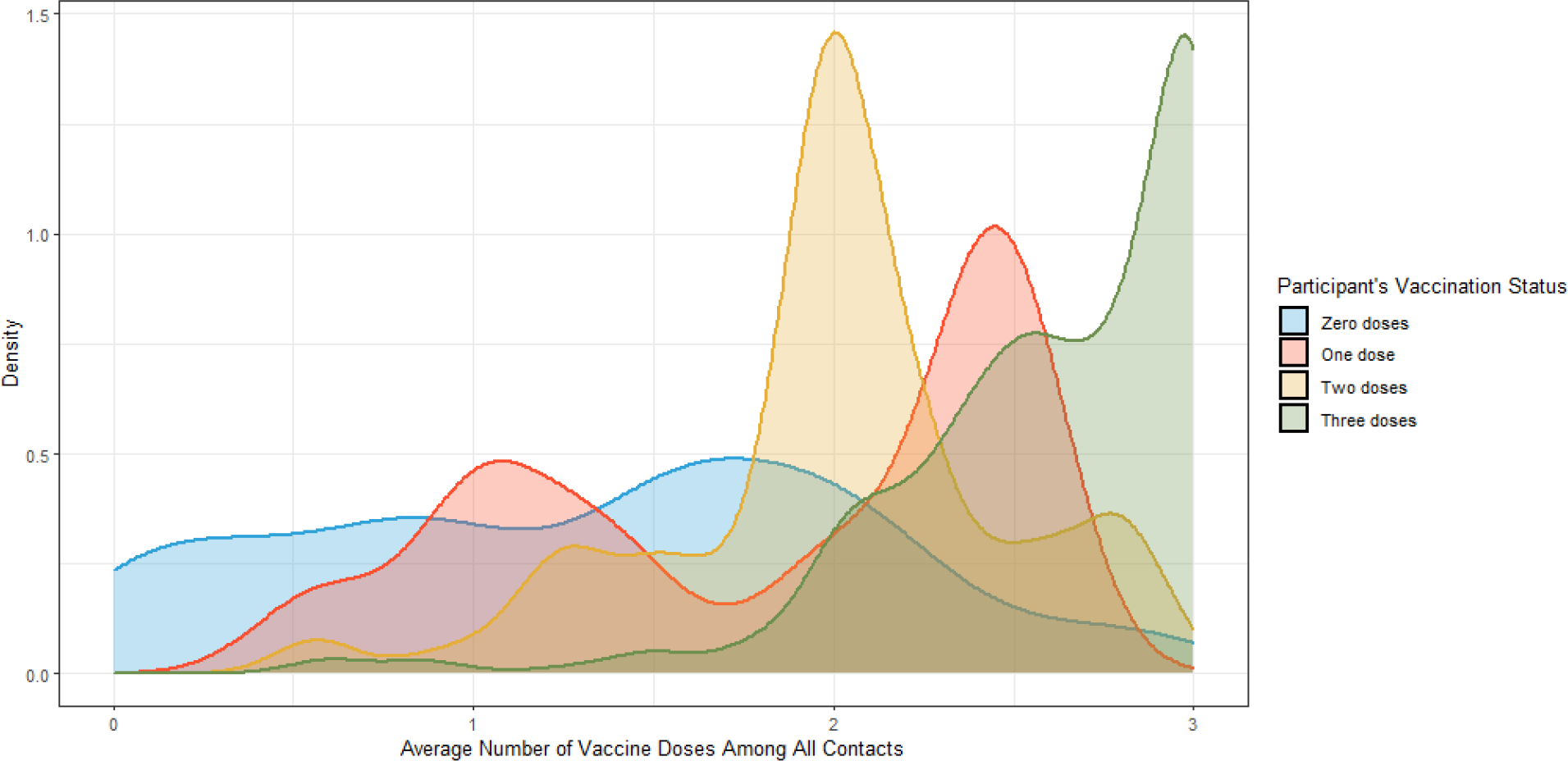
*Average Number of Doses Among All Contacts According to Self-Reported Vaccination Status.*

**Figure 3** shows the density curves for PAH scores stratified according to self-reported vaccination status. Based on the prevalence-adjusted vaccine homophily scores, all vaccination groups were more homophilous than would be expected based on the observed distribution of vaccination status (all *p*<0.05). In a simple regression model, those with three vaccine doses had significantly lower than expected homophily (*p*=0.007) compared to those with no doses and those with two doses had significantly higher homophily (*p*<0.001) than those with no doses. PAH was not significantly different between individuals with one dose and those with no doses (*p*=0.276). PAH, prevalence-adjusted homophily.

**Figure 3.**
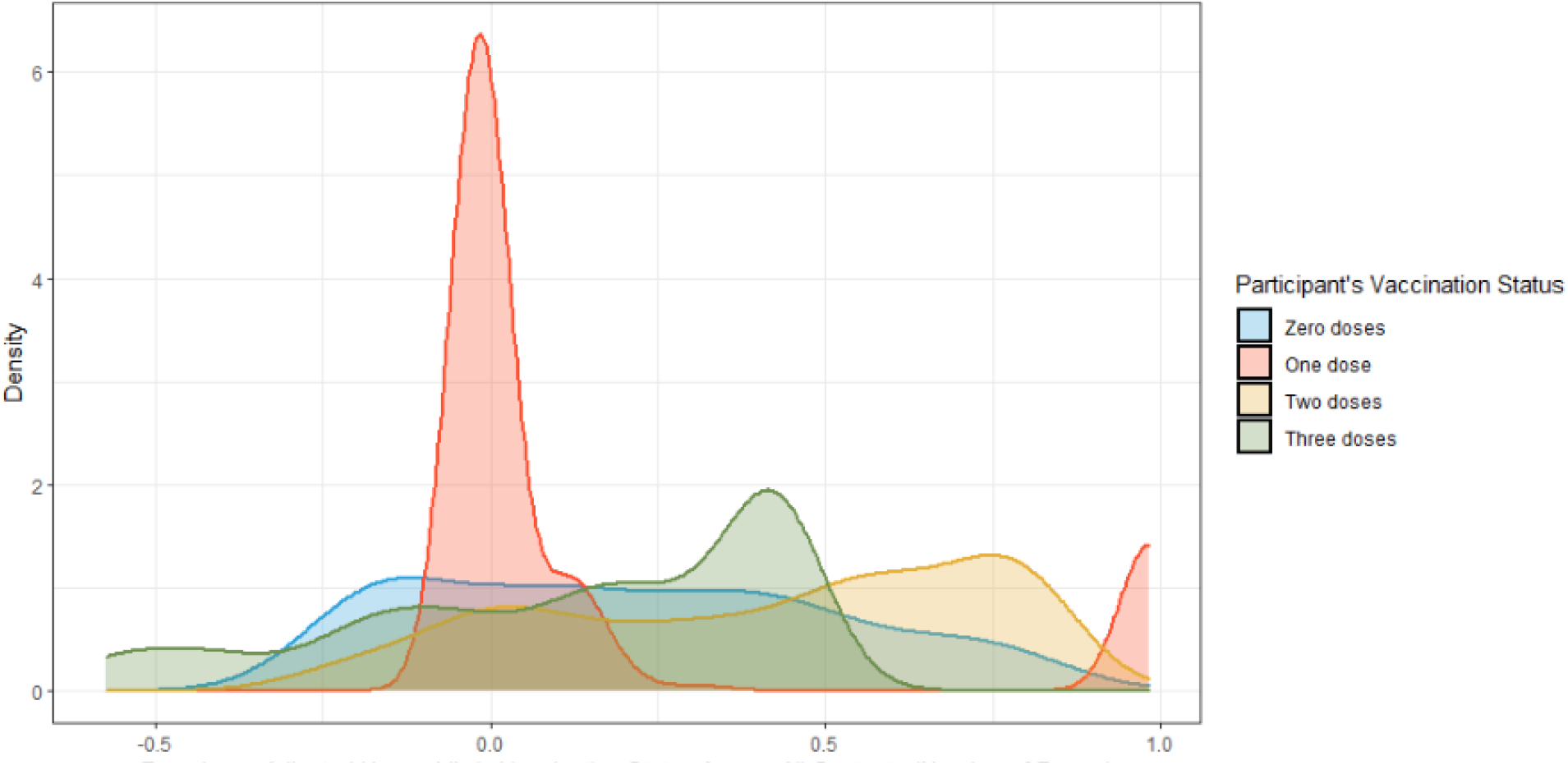
*Average Number of Doses Among All Contacts According to Self-Reported Vaccination Status.*

**Figure 4** shows boxplots of the participants’ contact network sizes stratified according to vaccination status. Participants with more vaccine doses—particularly those with three or more doses—tended to have smaller average network sizes (Spearman’s *r*=−0.217, *p*<0.0001).

**Figure 4.**
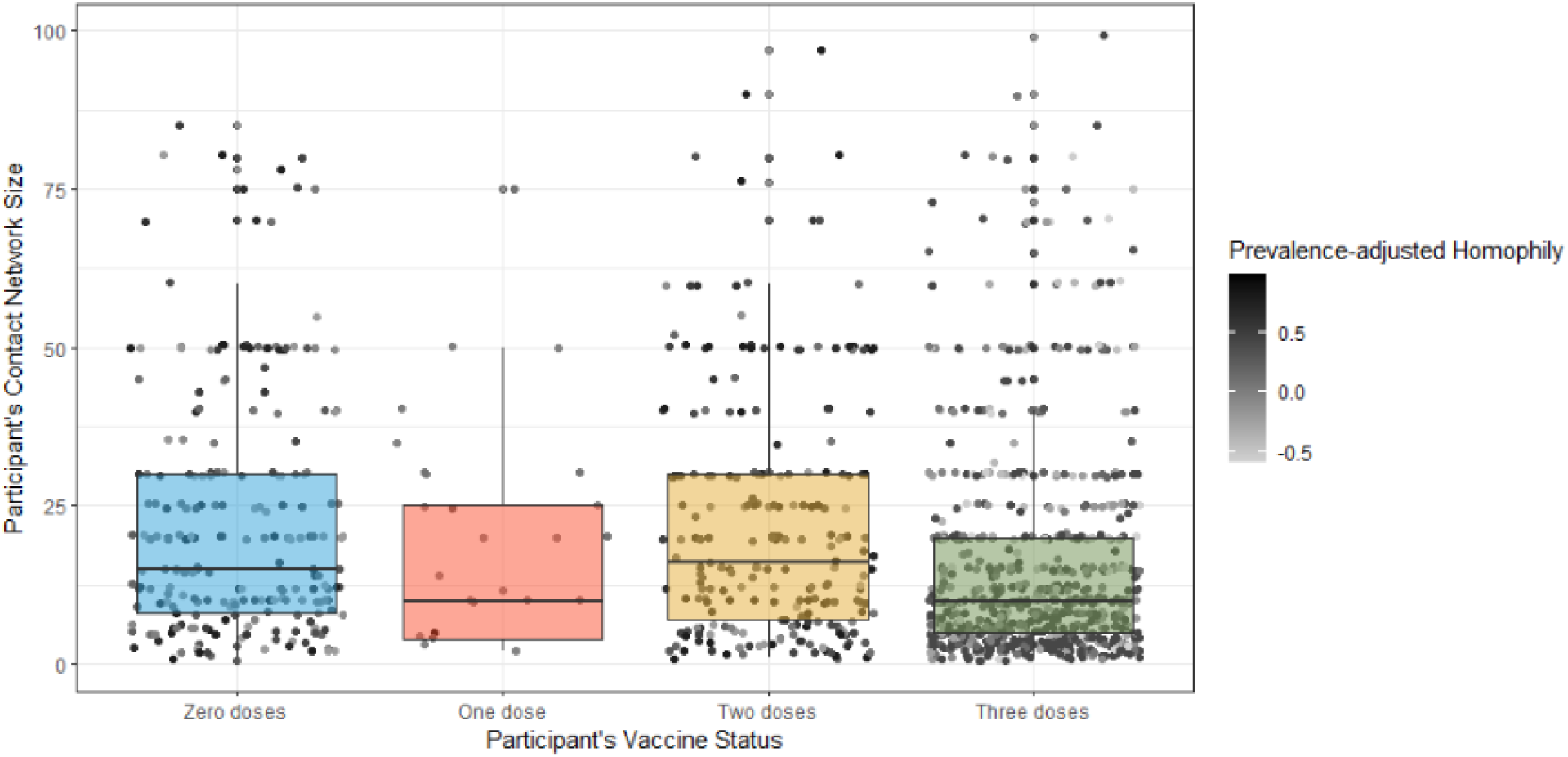
*Homophily and Contact Network Size by Quantile.*

### Aim 2. Demonstration of the Impact of Vaccine Homophily on COVID-19 Transmission

Our deterministic mathematical model tested the impact of vaccine homophily on COVID-19 transmission dynamics under conditions of high and low vaccine efficacy. To illustrate these effects, **Figure 5** presents four scenarios describing the intersection of vaccine homophily and vaccine efficacy. Each panel in the figure shows the number of infections from 0 to 60 days and two heat maps characterizing the force of infection at 15 (P1) and 45 (P2) days. Overall, in both low and high vaccine efficacy scenarios, the presence of vaccine homophily contributes to higher levels of epidemic growth. We describe each of the four scenarios in the following section to highlight the interaction between homophily and vaccine efficacy. The initial conditions were set up to reflect the vaccination uptake levels in British Columbia on February 16, 2023. The model was subsequently fitted to the case report data during the study period, and the last value of each state variable was used to initialize the model. The model was then simulated for two scenarios, each covering a 60-day period.

**Figure 5.**
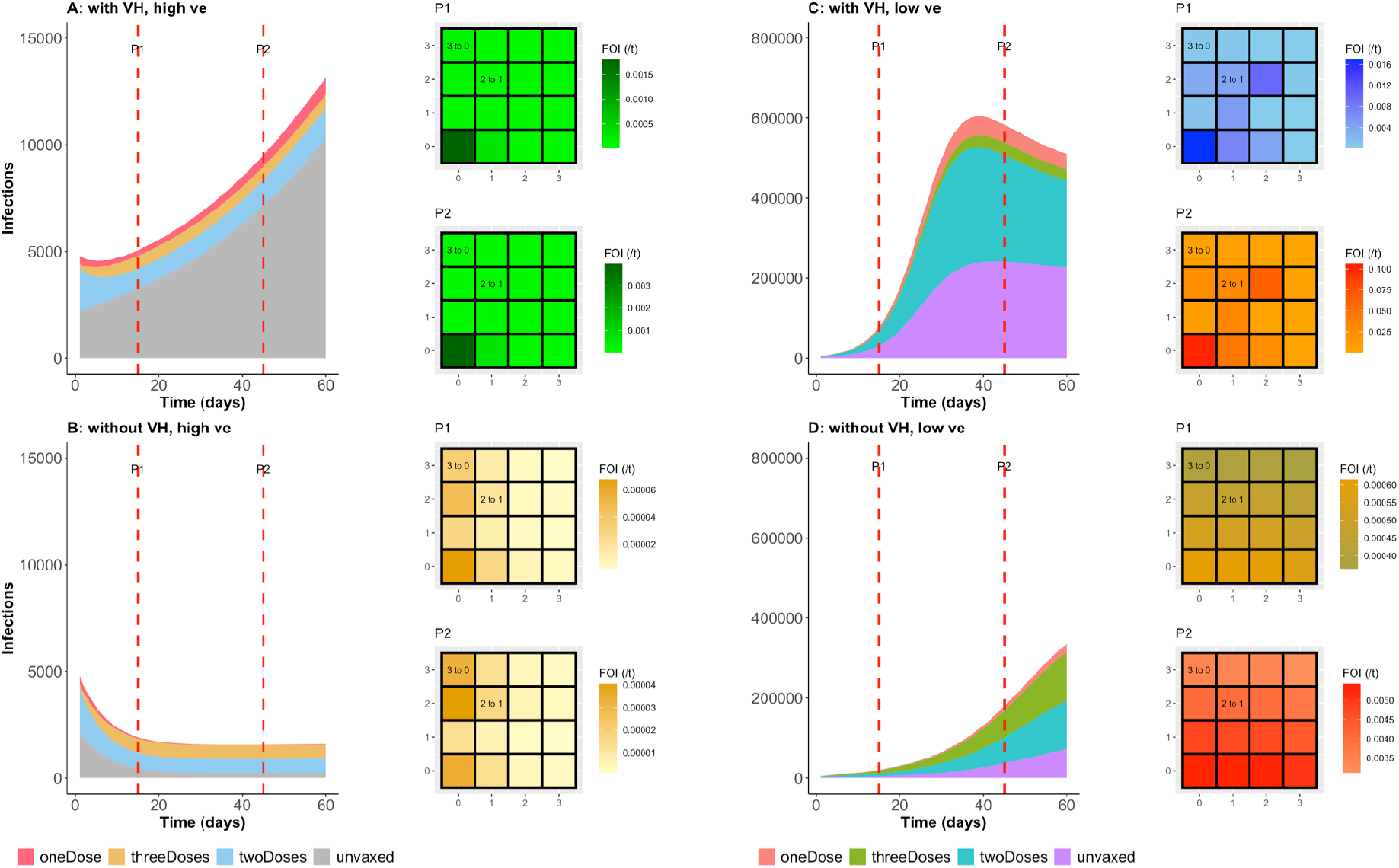
Number of Infections and Force of Infection: Assessment of the Impact of Homophily Under Scenarios of Low and High Vaccine Efficacy. *(A) Number of infections under a scenario with vaccine homophily and high vaccine efficacy. The trajectory is colour-coded by vaccination status. Heat maps P1 and P2 show the force of infection at 15 and 45 days, respectively. (B) Number of infections under a scenario without vaccine homophily and with high vaccine efficacy. Heat maps P1 and P2 show the force of infection at 15 and 45 days, respectively. (C) The number of infections per day for various vaccination groups under a scenario with vaccine homophily and low vaccine efficacy. Heat maps P1 and P2 show the force of infection at 15 and 45 days, respectively. (D) Daily number of infections according to vaccination status under a scenario without homophily and low vaccine efficacy. Heat maps P1 and P2 show the force of infection at 15 and 45 days, respectively. The following parameter values were used under conditions with vaccine homophily.* *p*_00_ = 0. 39, *p*_01_ = 0. 04, *p*_02_ = 0. 1552, *p*_03_ = 0. 414*p*_10_ = 0. 16, *p*_11_ = 0. 18, *p*_12_ = 0. 18, *p*_13_ = 0. 48, *p*_20_ = 0. 1293, *p*_21_ = 0. 0107, *p*_22_ = 0. 62, *p*_23_ = 0. 24, *p*_30_ = 0. 0462,, *p* _33_ = 0. 68, *p*_31_ = 0. 0038, *p*_32_ = 0. 27. 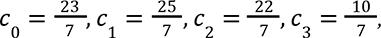 *l*_0_ = 1 − 0. 134, *l*_1_ = 1 − 0. 174, *l*_2_ = 1 − 0. 349, *l*_3_ = 1 − 0. 817*. Under conditions without vaccine homophily, we used: p_ij_* = 0. 25, 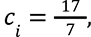 *l_i_* = 1 − 0. 514*. Under conditions of low vaccine efficacy, we used: v*_1_ = 1 − 0. 01, *v*_2_ = 1 − 0. 015, *v*_3_ = 1 − 0. 07, ν_0_ = 0. 2, ν_1_ = 0. 4, ν_2_ = 0. 65, ν_3_ = 0. 8*. Under conditions of high vaccine efficacy, we used: v*_1_ = 1 − 0. 6, *v*_2_ = 1 − 0. 89, *v*_3_ = 1 − 0. 93, ν_0_ = 0. 8, ν_1_ = 0. 85, ν_2_ = 0. 9, ν_3_ = 0. 97*. The following parameters were invariant:* 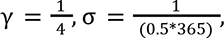 β = 0. 38, *f*_0_ = *f*_1_ = 0, *f*_2_ = *f*_3_ = 150 *per day. The horizontal and vertical axes on the heat maps represent vaccination status. The numbers within the heat maps indicate which group is transmitting infection to which other group: “2 to 1” indicates that individuals with 2 doses transmit to those with only 1 dose on that grid, and “3 to 0” indicates that those with ≥3 doses transmit to unvaccinated individuals on that grid*.

#### With vaccine homophily and high vaccine efficacy (Figure 5A)

In this scenario, the epidemic is driven and sustained primarily by unvaccinated individuals (see green heat maps), and even when infections decline initially in vaccinated groups, epidemic growth is sustained in the unvaccinated group. The force of infection under this scenario shows that transmission is sustained within the unvaccinated group with little effect on other groups, due to the high vaccine efficacy.

#### Without vaccine homophily and high vaccine efficacy (Figure 5B)

In this scenario, the epidemic declines faster in vaccinated individuals than in the unvaccinated group, while disease importation sustains transmission at a steady state. Moreover, some infections in the unvaccinated group are caused by the vaccinated groups due to the sizes of the groups and contact between them (see Figure S2 describing contact between groups). On the other hand, high vaccine efficacy against infection limits the force of infection from the unvaccinated group to the optimally vaccinated group.

#### With vaccine homophily and low vaccine efficacy (Figure 5C)

In this scenario, infections decline faster in the vaccinated groups than in the unvaccinated group, despite low vaccine efficacy. The epidemic is predominantly driven by unvaccinated individuals and those with two doses. This could be attributed to the large size of the ‘2-dose’ group combined with the relatively low vaccine efficacy. While each individual is partially protected, the overall population size and low vaccine efficacy result in the total force of infection from the ‘2-dose’ group being comparable to that from the unvaccinated population. Unvaccinated individuals greatly impact those within their group, and have some impact on those with one or two doses but minimal impact on the group with three or more doses, as mixing pattern limits intergroup contact. A similar pattern was observed in the group with two doses.

#### Without vaccine homophily and with low vaccine efficacy (Figure 5D)

In this scenario, each group affects itself and other groups equally, although the strength of the impact depends on the vaccination status. Moreover, the unvaccinated group has a disproportionate impact on both partially and optimally vaccinated groups.

#### Without vaccine homophily and with baseline scenario vaccine efficacy

In the baseline scenario (See Supplementary Material Figure S3), where parameter values are chosen to reflect vaccine efficacy against infection with the Omicron variant (34), the unvaccinated group drives infections in the one-dose and two-dose groups with some reduced impact on the group with three or more doses. However, the impact of the fully vaccinated group on the unvaccinated groups is less pronounced than the converse.

## Interpretation

### Primary Findings

This study was performed to characterize observable patterns of vaccine homophily and examine associations between vaccine homophily, self-reported vaccination status, COVID-19 prevention behaviours, contact network size, and self-reported COVID-19 infection. In addition, we examined the impact of vaccine homophily on COVID-19 transmission both within and between vaccination status groups under conditions of high and low vaccine efficacy. The results indicated the occurrence of vaccine homophily, with an average of 60% of the participants’ network contacts having the same number of vaccine doses as the participants themselves. Even adjusting for the population prevalence of each vaccine dose, each vaccination status showed higher than expected homophily. Similarly, the average number of doses received by household and non-household contacts was highest among those with ≥3 doses and lowest among those with 0 doses, demonstrating a higher prevalence of vaccination within the networks of vaccinated individuals relative to unvaccinated individuals. Those who were unvaccinated also had more diverse social networks with regard to vaccine status, were more likely to report previous COVID-19 infection, and had larger social network sizes. Mathematical models demonstrated that these dynamics contribute to elevated transmission overall under conditions of high vaccine efficacy, and transmission is driven primarily by unvaccinated individuals infecting other unvaccinated individuals. Under conditions of low vaccine efficacy, within-group transmission among unvaccinated individuals remains high, but there is also considerable impact of unvaccinated transmission on vaccinated individuals. Those with suboptimal protection (e.g., two doses) also experience considerable within-group transmission due to high contact rates with other suboptimally protected contacts within their network.

One factor contributing to these patterns is a higher level of observed vaccine homophily among household contacts compared to non-household contacts. Indeed, among unvaccinated participants, only 39% of household contacts had one or more doses of the COVID-19 vaccine, compared to 68% of non-household contacts. We also found that vaccine homophily appears to decrease as social network size increases, suggesting that tightly knit networks are more similar to one another than larger, distally connected networks. This is consistent with the empirical expectation that people tend to associate with people like themselves and are more different from those who are more socially distant (11).

To our knowledge, there have been few reports of empirically measured COVID-19 vaccine homophily. However, our findings that vaccine homophily has important implications for understanding the transmission of COVID-19 were consistent with previous modelling studies (12,13,15). In situating our findings within these previous studies, it is important to note that the impact of vaccine homophily differs according to the level of vaccine efficacy. Under conditions of high vaccine efficacy, transmission is largely among unvaccinated individuals, while contact patterns between groups put even fully vaccinated individuals at risk of infection under conditions of low vaccine efficacy. Further, contrary to some narratives that blame unvaccinated individuals for driving the epidemic under conditions of low vaccine efficacy, we found that the force of infection is substantially driven by contact networks and that a sizeable force of infection among unvaccinated individuals comes from those who are suboptimally vaccinated. With vaccine homophily, unvaccinated individuals pose significantly greater risk to other unvaccinated individuals than to other groups. The impact of unvaccinated individuals on fully vaccinated individuals is considerable only when there is low vaccine homophily and vaccine efficacy is low. For all the scenarios we considered, the impact of homophily is amplified by increased probability of infection per contact.

The overrepresentation of the unvaccinated in the total number of infections (Fig 2A) is similar to findings from Canada, based on case-level vaccine history data. Among individuals aged 5 years and older, the unvaccinated constitute 41% of the 73% of total reported cases since the onset of the vaccination rollout, as of June 10, 2022. As the vaccination rollout progresses, the limited testing capacity has resulted in the targeted testing of the high-risk population for severe disease, which coincides with the group prioritized during the vaccination rollout. Consequently, this bias in the case report data indicates that reported case data by vaccination status may not accurately reflect the distribution of infections by vaccination status at the population level (35). For example, in British Columbia, the unadjusted data indicated that the unvaccinated accounted for 14.2% of the total cases, whereas the age-adjusted cases per 100,000 population in the province showed that unvaccinated groups accounted for 58% in March 2022 (36). This finding is consistent with the initial conditions of our model at the beginning of March 2022.

Taken together, our findings are worrisome, particularly when considering the risk for transmission within households, which are known to account for a significant proportion of COVID-19 infections (37-39). Further, given the clustering of risk among unvaccinated individuals even when an effective vaccine is available has important implications for considering vaccine-status-specific COVID-19 prevention measures, such as mask mandates, physical distancing rules, and proof of vaccination requirements. Given the group transmission dynamics that arise due to household and non-household contact networks, it is important to engage these populations to address vaccine hesitancy (40-44). This will likely require community-based and culturally aware public health interventions that can help reduce vaccine hesitancy. Indeed, rather than viewing unvaccinated individuals as a threat to public health, it should be taken as an opportunity to educate and work with these individuals to address their concerns, particularly given the skepticism that may be associated with the emergency use authorizations that have allowed the rapid rollout of COVID-19 vaccines (43,44).

## Limitations

This study had some limitations that should be taken into consideration when interpreting our findings. First, we note that our findings are relevant to the promotion of vaccines across the population and emphasize the importance of continued vaccine research and efforts to provide ongoing protection as vaccine-induced immunity wanes. However, our data are from a period in which individuals were receiving third doses and facing the rising prevalence of the Omicron variant. Therefore, our results should not be read as predictive scenarios. Rather, they should be interpreted in the context of a pandemic-related mass-vaccination effort, during which there was uneven uptake of vaccines across social networks due to a variety of factors within and outside the control of individuals. Second, we note that our survey utilized an online, opt-in convenience sampling methodology to study the effects of interest. Online sampling is now a widespread methodology, particularly since the decline in reliability of other opt-in sampling strategies such as random-digit dialing methods. Point estimates from this study are therefore likely to be non-representative and may be biased. However, we note that studies show that epidemiological and behaivoural estimates from web and telephone surveys are typically comparable, and that online samples may have advantages to other survey methods (e.g., reduced favourable reporting;47). This is because the direction of bias may be random While population weights may partially adjust for this issue, the direction and magnitude of potential biases are unknown. Replication in a population-based sample is warranted. Third, it is important to acknowledge that our sample size was relatively modest. Replicating our findings in a larger sample could offer more robust evidence and enhance the accuracy of our measurements. However, we must acknowledge that replicating the study will present significant challenges, particularly given the current stage of the pandemic. Tracking the vaccination statuses of individuals within contact networks may prove to be a daunting task. Fourth, self-reported data may be unreliable, particularly estimates regarding characteristics of participants’ social networks. People may be overly confident in estimating their vaccination status, guideline compliance, and vaccine history of their social network contacts, which may result in a systematic bias toward the hypothesis that vaccine status homophily exists. Fifth, we do not intend to imply causality in describing any of the relationships between vaccine status and vaccine homophily. Further qualitative and quantitative studies are needed to understand the processes that give rise to vaccine homophily and how best to respond to these network characteristics.

## Conclusion

The present study identified evidence of homophily in COVID-19 vaccine uptake. Unvaccinated individuals are more likely to have unvaccinated network contacts, conditions that create increased risk of COVID-19 transmission among unvaccinated individuals. Nevertheless, vaccine homophily varies considerably, and further research is needed to understand the factors that shape vaccine homophily within social networks. Vaccine status-specific prevention guidelines may help to mitigate the risks to communities posed by the unique risk profiles of unvaccinated individuals.

## Data Availability

Data sets generated during the current study are available from the authors on reasonable request

## Declarations

### Ethics approval

The study protocol was reviewed by the Research Ethics Board at Simon Fraser University (Protocol #30000753)

### Consent to participate

All participants gave consent for participation

### Availability of data and material

Available on request

### Data availability

Data sets generated during the current study are available from the authors on reasonable request

### Author contributions

Study design (KC, CC), Data collection (KC), Initial data analysis (KC, CC), Model conceptualization (EBA, CC), Simulation (EBA), Initial Draft (KC), Visualization (EBA, KC), Funding acquisition (KC, CC), Supervision (CC), Resources (KC, CC) Writing, Editing and Proofreading (EBA, KC, CC)

## Supplementary Information

**Table S1.** Variables

| **Variable** | **Measurement** |
| --- | --- |
| COVID-19 infection history | “Have you ever been diagnosed with COVID-19?” Participants could choose from the following responses:<br>(a) Yes, I have been diagnosed with COVID-19<br>(b) No, but I think I have had COVID-19. I just never received a test and/or diagnosis<br>(c) No, and I do not think I have had COVID-19 |
| Compliance with provincial mandates and guidelines | “On a scale of 1 (Not At All) to 4 (Very Closely), how closely have you followed provincial mandates and guidelines for COVID-19 prevention?” Participants could choose from the following responses:<br>(a) Not at all<br>(b) Not very closely<br>(c) Somewhat closely<br>(d) Very closely |
| Vaccination status | “How many doses of the COVID-19 vaccine have you received so far?” Participants could choose from the following responses:<br>(a) 0 doses<br>(b) 1 dose<br>(c) 2 doses<br>(d) 3 or more doses |
| Perceived COVID-19 history of regular contacts | “Thinking about the people you have regular contact with, how many of them have had COVID-19?” They were given the further instruction, “If you are unsure about how many have had COVID-19, provide your best estimate.” Participants could choose from the following responses:<br>(a) A few (i.e., 0%–20%) |
|  | (b) Some of them (i.e., 21%–40%)<br>(c) Around half of them (i.e., 41%–60%)<br>(d) Most of them (i.e., 61%–80%)<br>(e) Nearly all of them (i.e., 80%–100%) |
| Perceived compliance of regular contacts with COVID-19 prevention guidelines and mandates | “Thinking about the people you have regular contact with, how many of them have followed provincial guidelines for COVID-19 prevention ‘very closely’?”<br>(a) A few of them (i.e., 0%–20%)<br>(b) Some of them (i.e., 21%–40%)<br>(c) Around half of them (i.e., 41%–60%)<br>(d) Most of them (i.e., 61%–80%)<br>(e) Nearly all of them (i.e., 80%–100%)<br>They were given the further instruction, “If you are unsure about whether others are following guidelines ‘very closely,’ provide your best estimate.”<br>“If you had to provide an exact percentage, what percentage of those you have regular contact with are vaccinated?” |
| Vaccination status of household contacts | “Excluding yourself, how many people 12 years of age or older live in your household?”<br>The following clarification was also provided: “By household we mean anyone living at the same address as you, that you share a kitchen with.” |
| Number of people in their household that had received each level of vaccine dose | “For each person in your household, aged 12 years or older and excluding yourself, how many belong to each category.” The following clarification was also provided: “The total for the four categories below should sum to <<household size provided above>>, which you indicated was the number of people living in your household, excluding yourself.”<br>Participants could choose from the following |
|  | responses:<br>(a) 0 doses.<br>(b) 1 dose.<br>(c) 2 doses.<br>(d) 3 or more doses.<br>(e) Unknown |
| Vaccination status of non-household contacts | “The following questions are about the vaccination status of people you have been in contact with over the past 7 days, excluding members of your household. Please keep in mind the following as you answer these questions: COVID-19 is an airborne respiratory disease spread through saliva, other bodily fluids, and aerosols (such as those exhaled when talking, singing, sneezing, coughing, or breathing). Face-to-face contact, direct physical contact, and sharing the air of a person with COVID-19 in a poorly-ventilated indoor space increase your exposure to coronavirus-containing aerosols.” |
| Number of contacts | “In the past 7 days, how many people, regardless of whether they had COVID-19 or not, have you had contact with?” The following clarification was also provided: “Please do not include members of your household in this count. In providing this number, please use your best estimate. Please, include only people whom you interacted with directly.” |
| Number of contacts for whom they knew the vaccination status | “In the previous question, you reported that you had contact with <<*non-household contact network size provided above*>> people in the past 7 days. Of these people, how many do you know the vaccination status of?” Participants were reminded that, “This number should be the same or smaller than the number (i.e., <<*non-household contact network size provided above*>>) that you provided in the previous question. In the next question we will ask you to report on the |
|  | vaccination status for these people.” |
| Number of people among their non-household contacts that had received each level of vaccine dose | , “For people who you were exposed to in the past 7 days, excluding members of your household, how many belong to each category? The following clarification was also provided: “The total for the four categories below should sum to <<*contacts whom participants knew the status of*>>, which you indicated was the number of people who you had been in contact with and whose vaccination status you knew.”<br>Participants could choose from the following responses:<br>(a) 0 doses.<br>(b) 1 dose.<br>(c) 2 doses.<br>(d) 3 or more doses.<br>(e) Unknown |

**Figure S1.**
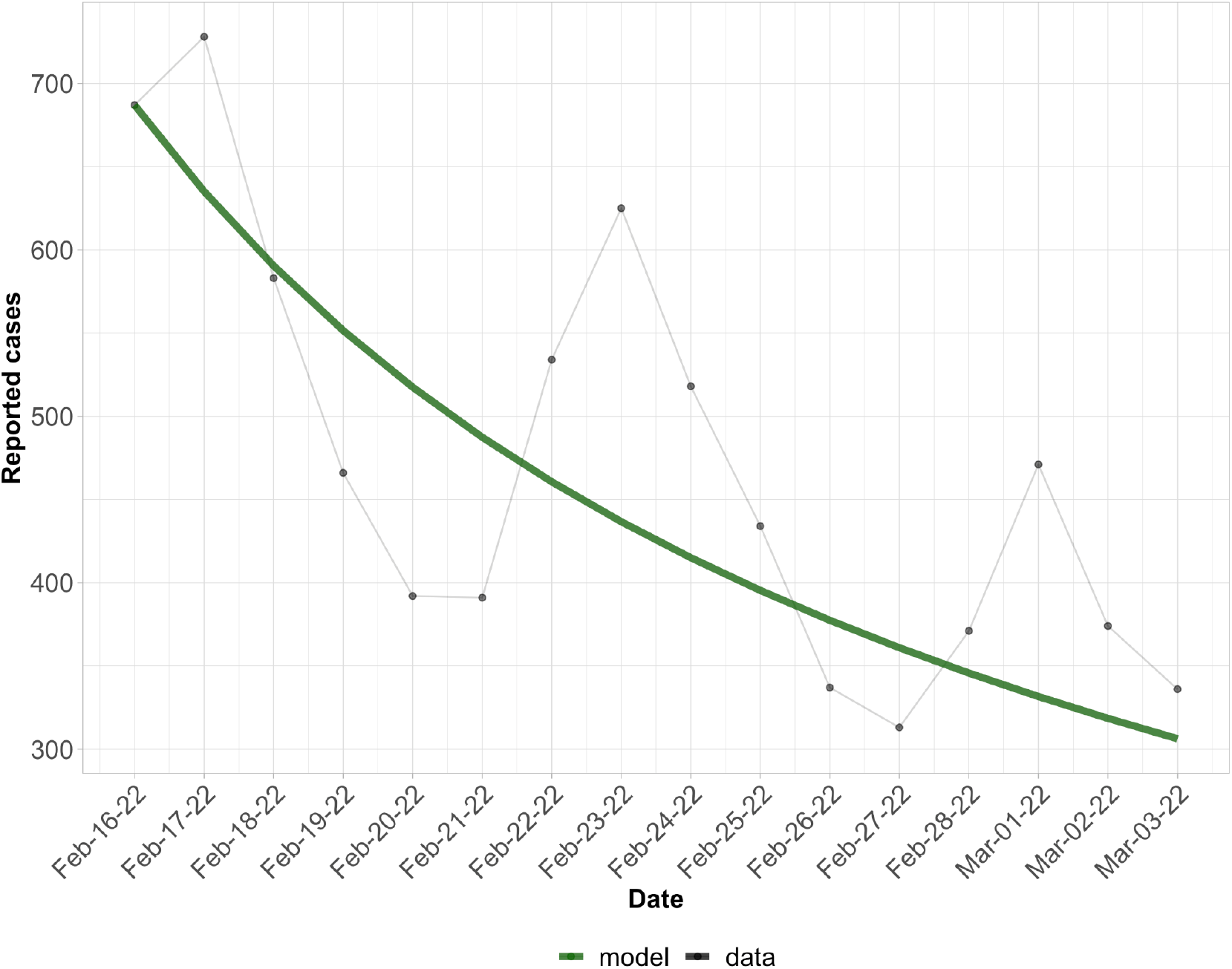
*Model Fit to Reported Cases From February 16, to March 3, 2022.* *The green line indicates the model output, while the gray connected dots show reported cases during the study period*.

**Figure S2.**
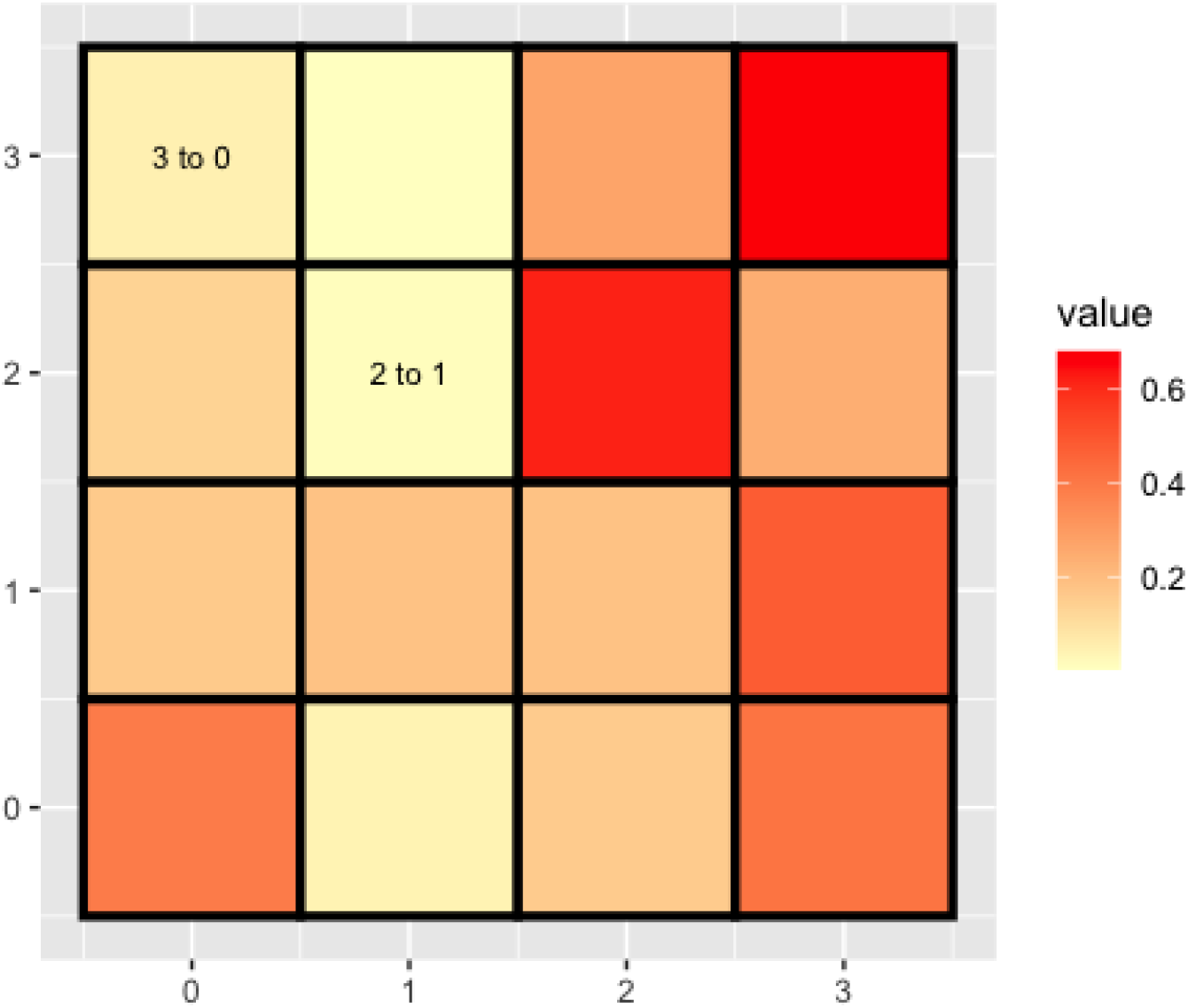
*A Null Model Showing the Proportions of Contacts of Each Vaccination Group.* *Each panel shows the proportion of contacts either within or outside their vaccination group. “2 to 1” indicates the proportion of contacts of individuals in the 2-dose group that have had 1 dose of vaccine. Axis labels indicate vaccination status*.

**Figure S3.**
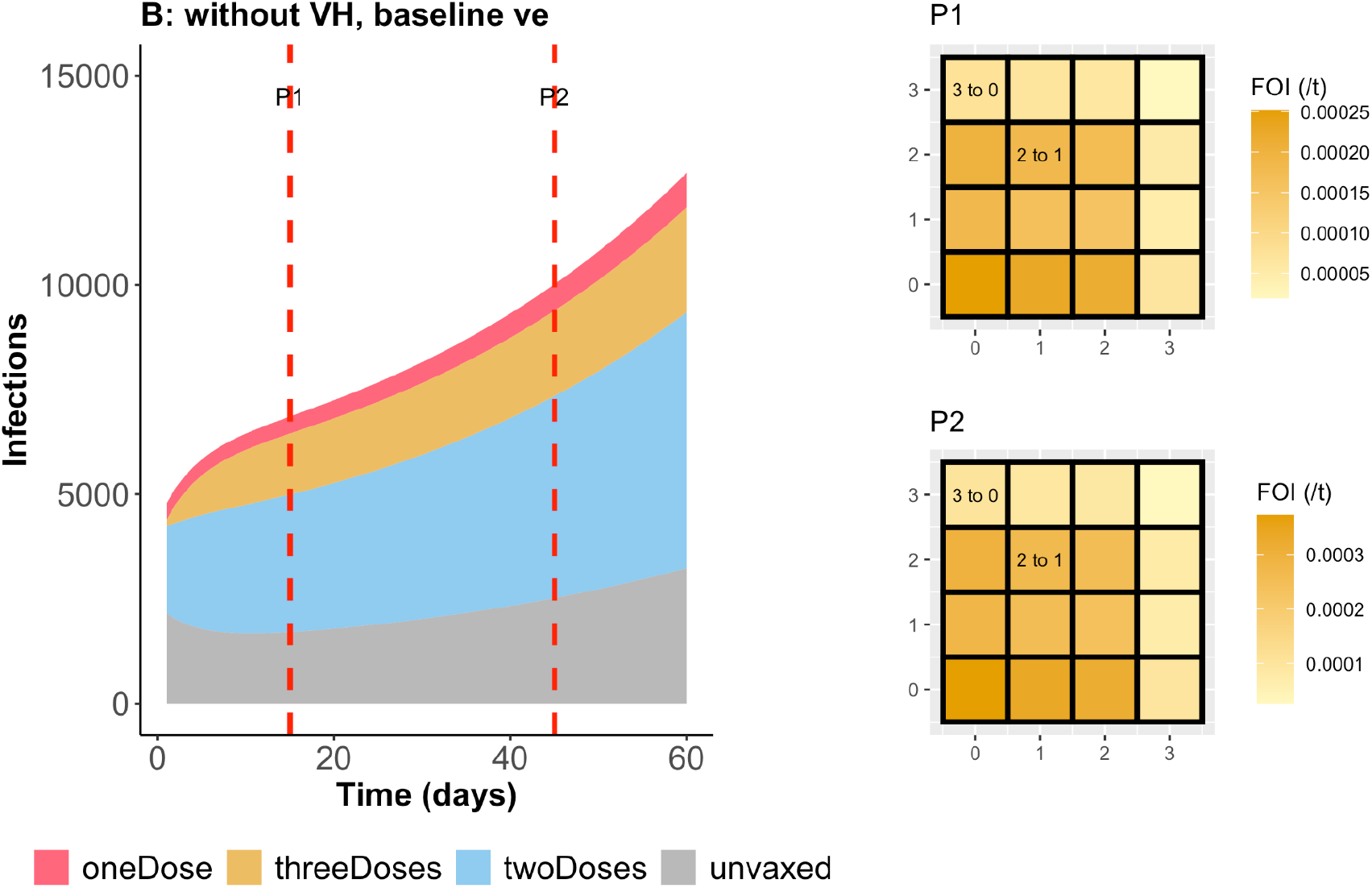
Baseline Scenario Showing Infections by Vaccination Status and Force of Infection Generated by Vaccination Groups on Days 15 and 45. The baseline parameter values are as follows (other parameters take the same values as in Figure 5 in the main text unless otherwise noted): *v*_0_=0.35,*v*_1_=0.65, *v*_2_=0.68,*v*_3_=0.83 and *v*_1_ = 1 − 0. 1, *v*_2_ = 1 − 0. 148, *v*_3_ = 1 − 0. 74.

